# The impact of COVID passport mandates on the number of cases of and hospitalizations with COVID-19 in the UK: a difference-in-differences analysis

**DOI:** 10.1101/2022.02.24.22271325

**Authors:** Kim López-Güell, Albert Prats-Uribe, Martí Català, Clara Prats, Jotun Hein, Daniel Prieto-Alhambra

## Abstract

**Background:** Mandatory COVID-19 certification was introduced at different times in the four countries of the UK. We aimed to study the effect of this intervention on the incidence of cases and hospital admissions.

**Methods:** The main outcome was the weekly averaged incidence of COVID-19 confirmed cases and hospital admissions. We performed Negative Binomial Segmented Regression (NBSR) and Autoregressive Integrated Moving Average (ARIMA) analyses for the four countries (England, Northern Ireland, Scotland and Wales), and fitted Difference-in-Differences (DiD) models to compare the latter three to England, where COVID-19 certification was imposed the latest.

**Findings:** NBSR methods suggested COVID-19 certification led to a decrease in the incidence of cases in Northern Ireland, but not in hospitalizations. In Wales, they also caused a decrease in the incidence of cases but not in hospital admissions. In Scotland, we observed a decrease in both cases and admissions. ARIMA models confirmed these results. The DiD model showed that the intervention decreased the incidence of COVID compared to England in all countries except Wales, in October. Then, the incidence rate of cases already had a decreasing tendency, as well as in England, hence a particular impact of Covid Passport was less obvious. In Wales, the model coefficients were 2.2 (95% CI -6.24,10.70) for cases and -0.144 (95% CI -0.248, -0.039) for admissions in October and -7.75 (95% CI -13.1, -2.46) for cases and -0.169 (95% CI-0.308, -0.031) for admissions in November. In Northern Ireland, -10.1 (95% CI -18.4, -1.79) for cases and -0.269 (95% CI -0.385, -0.153) for admissions. In Scotland they were 7.91 (95% CI 4.46,11.4) for cases and -0.097 (95% CI - 0.219,0.024) for admissions.

**Interpretation:** The introduction of mandatory certificates decreased cases in all countries except in England. Differences on concomitant measures, on vaccination uptake or Omicron variant prevalence could explain this discrepancy.

## Introduction

More than a year after the emergence of SARS-CoV-2, widespread transmission is arguably higher than ever. To date, the virus has caused more than 17,750,000 confirmed cases, 700,000 hospital admissions and 178,000 deaths in the UK^1^. All around western countries there has been a need to balance restrictions to fight the pandemic while controlling their impact on society.

Soon after the arrival of the Delta variant of the virus to the UK over the spring, it became the dominant one. Recently, the Omicron variant entered the UK, its first cases being reported in late November. The UK Health Security Agency estimates the current prevalence of Omicron to be higher than 90% as of end-January, having quickly overcome Delta as the most common variant^2^.

Since the emergence of the virus, various non-pharmaceutical interventions were introduced by several countries in Europe to fight against the COVID-19 pandemic, with adverse socio-economic effects. Their aim was to slow down the transmission by restricting mobility and social interactions, e.g. mass gathering measures. Several papers suggest some of them had an effect in reducing COVID-19 transmission^3–5^. Recently, mandatory COVID-19 certification regulating access to public venues, nightclubs or cultural events was implemented in some countries, using proof of at least two doses of an approved vaccine, negative test (usually within the last 2 days) or a recovery certificate of a recent infection (usually within the previous 6 months)^6^. Many voices have expressed concerns over its effectiveness and due to their potentially negative effects on the economy e.g. on the hospitality sector. Some studies report increased vaccine uptake after its implementation^7,8^, but there is a lack of research on its potential impact in reducing incidence of COVID-19.

The UK implemented COVID-19 certification (Covid Passport) during the second half of 2021, and each of its countries did it at different times. We took advantage of this natural experiment to study whether Covid Passport in the UK had an effect in reducing the incidence of COVID-19 cases and hospitalizations, considering the four countries separately. We use England as a negative control, since it was the last country where Covid Passport was introduced.

## Methods

### Data

Data on COVID-19 cases and hospital admissions in the UK was gathered from the UK Coronavirus Dashboard^1^, which is updated every day. Data on the implementation of the Covid Passport in Scotland, England, Northern Ireland and Wales was collected from official sources, as mentioned in media^9–12^. For all sources, we used data from the 1^st^ of January 2021 to the 19th of January 2022. Data was extracted on the 19th of January 2022. Data from mid-year 2021 population for each country was extracted from the UK Coronavirus Dashboard^1^ as well.

### Covid Passport

We studied the four countries of the UK (England, Scotland, Northern Ireland and Wales). A country was considered as implementing Covid Passport (CP) if the certificate was required for at least some frequently used public venues such as restaurants, nightclubs or cultural events. Scotland implemented Covid Passport on the 18^th^ of October, Northern Ireland did it on the 29^th^ of November. In the case of Wales, we modelled two different changes in the restriction of the certificate, as Covid Passport was first implemented for nightclubs on the 11^th^ October 2021 and then extended to cinemas, theatres and concert halls on the 15^th^ November 2021. England was the last country to require the certificate, only doing so after the 15^th^ of December. See *Supplementary material* for further detail on each country’s implementation of the Covid Passport.

### Outcomes

We studied two outcomes, for which we assessed the effect of the Covid Passport intervention: incidence rate of COVID-19 confirmed cases and incidence rate of COVID-19 hospital admissions in the general population. We introduced a lag after it to neglect data right after the intervention date, for which its effects were not expected to be significant. The lag was set to 5 days for COVID-19 cases and to 7 days for COVID-19 hospital admissions^13^.

### Study time intervals

We selected the time intervals for the study of each intervention as wide as possible, provided that they did not include more than one change in the intervention, that they included more than 10 points (days) at each side of the lag interval and that they did not show, if possible, exogenous changes in convexity.

### Statistical analysis

We calculated incidence rates as number of cases (COVID-19 or admissions) divided per each country’s population. We also calculated 7-day smoothed rolling average rates to reduce the effects of lower reporting on weekends.

We performed the first analysis on the 7-day smoothed data using Negative Binomial Segmented Regression (NBSR). We preferred this method to a linear model as it provides a more accurate representation of count data, and also allows for over-dispersion. We selected the time point of the intervention as specified before. The method provides insight on both changes in level and trend of the variable of interest after the intervention, and also allows for predictions of outcomes had the intervention not been put into effect.

As NBSR models cannot account for auto-correlation of the outcome, we also fitted Autoregressive Integrated Moving Average (ARIMA) models to the same time frames to estimate the effect of the interventions while controlling for potential association of the observations. In NBSR we are interested in studying the regression coefficients, of which a negative value is indicative of a reduction effect of the variable on the response. The exponential of the coefficients is used to extract the changes in slope directly. We assess both a step change and in the slope before and after the intervention. Note that, while the former indicates a local reduction, the slope change is much more interesting and accounts for a temporarily longer effect. Even though ARIMA models do not allow for such a simple interpretation of coefficients, their sign can be broadly considered in the same way, while controlling for autocorrelation.

To further strengthen the results, and given that England did not implement the Covid Passport when it was effective in the other three countries, we used its data as a counterfactual for Difference-in-Differences (DiD) models. These methods compare the mean of the variable of interest for an exposed and control groups and before and after of a certain interrupting point, providing hence insight on the changes of the variable for the exposed countries relative to the change in the negative outcome group. We cannot draw causal conclusions by simply observing before-and-after changes in outcomes, because other factors might influence the outcome over time, and DiD methods overcome that by introducing a comparison between two similar groups exposed to different conditions. First, DiD takes the difference for both groups before and after the intervention. Then it subtracts the difference of the control group to the exposed one to control for time varying factors, thus estimating the clean impact of the intervention. In essence, the DiD estimating equation is the following:

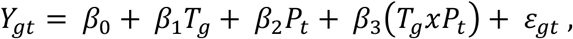

where *Y*_*gt*_ is the outcome for an individual in group g and treated unit t, *P*_*t*_ is a binary time variable indicating whether the observation belongs to the period before or after the intervention and *T*_*g*_ is a binary variable indicating whether the observation belongs to the exposed or the controlled group. In this setting, the treatment effect is estimated with the coefficient *β*_3_ from the regression.

We also considered the necessary conditions for the comparison to be sensible, namely all the assumptions of the OLS model and the parallel trends assumption, which requires both groups to present similar trends before the intervention time point.

We performed all the analyses in R v4.3 and used the packages epiR, tidyverse, forecast, ggplot2, MASS and lmtest. Code is available in https://github.com/KimLopezGuell/Covid-passport.

## Results

*Table 1* provides estimates and 95% confidence intervals for the slope (t1) before the intervention, the step change of the non-pharmaceutical intervention (NPI) after the lag time and the change in the slope (t2) after the lag time for all NBSR models. The results suggested that Covid Passport was associated with the reduction of cases for all the countries of the UK apart from England, as well as with the reduction of hospital admissions for Scotland. The results from NBSR were reinforced by the respective ARIMA models.

**Table 1:** Estimates and 95% confidence intervals of the effect of the Covid Passport in the NBSR and ARIMA models for the different countries and outcomes. NBSR coefficients are exponentiated. Information on trend pre-intervention (t1) and trend (t2) and level (NPI) change estimates after intervention are provided. *CP+number* indicates the number-th Covid Passport intervention.

|  |  |  | Wales CP1 | Wales CP2 | Northern<br>Ireland CP1 | Scotland CP1 | England<br>CP1 |
| --- | --- | --- | --- | --- | --- | --- | --- |
| Cases | NBSR | Estimate t1 (%) | 1.01 | 1.01 | 1.01 | 1.01 | 1.01 |
|  |  | 95% CI t1 (%) | (0.99,1.04) | (0.99,1.04) | (1.00,1.02) | (0.97,1.04) | (1.00,1.02) |
|  |  | Estimate t2 (%) | 0.97 | 0.98 | 0.99 | 0.99 | 1.02 |
|  |  | 95% CI t2 (%) | (0.94,0.99) | (0.95,1.02) | (0.97,1.01) | (0.95,1.03) | (1.00,1.04) |
|  |  | Estimate NPI (%) | 1.25 | 0.82 | 0.87 | 0.96 | 1.76 |
|  |  | 95% CI NPI (%) | (0.98,1.59) | (0.61,1.12) | (0.73,1.04) | (0.67,1.40) | (1.54,2.00) |
|  | ARIMA | Estimate t1 (%) | -0.33 | 1.15 | 1.05 | 0.33 | 1.31 |
|  |  | 95% CI t1 (%) | (-3.03,2.38) | (-0.65,3.06) | (0.55,1.55) | (-2.70,3.37) | (0.39,2.23) |
|  |  | Estimate t2 (%) | -1.73 | -1.62 | -0.16 | -2.30 | 4.07 |
|  |  | 95% CI t2 (%) | (-4.61, 1.15) | (-4.35, 1.11) | (-3.05, 2.73) | (-5.46, 0.87) | (1.48, 6.67) |
|  |  | Estimate NPI (%) | 39.30 | -10.60 | -12.40 | 32.33 | 78.00 |
|  |  | 95% CI NPI (%) | (7.85, 70.79) | (-31.31, 10.13) | (-30.25, 5.43) | (-0.37, 65.03) | (44.93, 111.04) |
| Admissions | NBSR | Estimate t1 (%) | 1.00 | 0.99 | 0.98 | 1.01 | 1.00 |
|  |  | 95% CI t1 (%) | (0.81,1.24) | (0.92,1.06) | (0.94,1.02) | (0.86,1.19) | (0.97,1.04) |
|  |  | Estimate t2 (%) | 1.00 | 1.00 | 1.02 | 0.97 | 1.01 |
|  |  | 95% CI t2 (%) | (0.77,1.31) | (0.89,1.12) | (0.91,1.13) | (0.77,1.22) | (0.96,1.06) |
|  |  | Estimate NPI (%) | 1.22 | 0.89 | 1.17 | 0.96 | 1.89 |
|  |  | 95% CI NPI (%) | (0.09,22.29) | (0.19,4.36) | (0.31,4.19) | (0.12,8.53) | (0.74,5.12) |
|  | ARIMA | Estimate t1 (%) | 0 | -0.02 | -0.03 | -0.67 | 0 |
|  |  | 95% CI t1 (%) | (-0.05,0.04) | (-0.03,0) | (-0.04,0) | (-4.07,2.74) | (-0.01,0.03) |
|  |  | Estimate t2 (%) | 0.02 | 0.02 | 0.01 | -2.16 | 0.05 |
|  |  | 95% CI t2 (%) | (-0.04, 0.07) | (-0.01, 0.04) | (-0.03, 0.06) | (-6.98, 2.65) | (0.02,0.09) |
|  |  | Estimate NPI (%) | 0.32 | -0.11 | 0.35 | 34.33 | 1.14 |
|  |  | 95% CI NPI (%) | (-0.25, 0.89) | (-0.39, 0.17) | (-0.19, 0.88) | (-11.20, 79.87) | (0.53,1.75) |

*Supplementary Figure S1* suggests that the introduction of Covid Passport stopped the increase in incidence rate of cases observed in early December 2021 in Northern Ireland, while it peaked in England, where no Covid Passport restriction was introduced, during the same time. We observed a daily raise of 1.01 % (95% CI 1.00,1.02) before the introduction of Covid Passport, which changed the slope to 0.99% (95% CI 0.97,1.01) afterwards. As for incidence rate of hospital admissions, Northern Ireland shows a decreasing trend during November, with a daily increase of 0.98% (95% CI 0.94,1.02), similar to England. Yet while in the latter the incidence rate increased rapidly with the arrival of the Omicron variant in the UK, the increase in Northern Ireland was much more moderate. With the introduction of the Covid Passport in the country, the slope of the trend increased to 1.02% (95% CI 0.91,1.13).

Regarding Scotland, in *Supplementary Figure S2* we note that the incidence rate of cases presented an initial increase of 1.01% per day (95% CI 0.97,1.04), which decreased to 0.99% (95% CI 0.95,1.03) after the intervention. As for the incidence rate of hospital admissions, the initial increase was of 1.01% per day (95% CI 0.86,1.19) and we observed a slope after Covid Passport introduction of 0.97% (95% CI 0.77,1.22).

While Covid Passport was associated with a slowdown of the incidence of cases in Wales during the second half of November, that was not seen in England (*Supplementary Figure S3a*). For Wales the pre-intervention slope was 1.01% (95% CI 0.99,1.04), which decreased to 0.97% after the intervention (95% CI 0.94,0.99). Likewise, hospital admissions (*Supplementary Figure S3b*) increased in England while decreasing in Wales in the same period.

An assessment of England (*Supplementary Figure S4*) itself during its introduction of Covid Passport on the 15^th^ December 2021 provided an estimate of the increase pre-intervention of 1.01% (95% CI 1.00,1.02) and a change in the slope to 1.02% (95% CI 1.00,1.04) for the incidence rate of cases. As for hospital admissions, the increase before the intervention was of 1.00% (95% CI 0.97,1.04) which changed to 1.01% (95% CI -0.96,1.06).

*Table 2* contains results of a DiD regression for both cases and admissions incidence rates of the different UK countries (Wales, Northern Ireland, Scotland) compared to England. Except from cases in Scotland and cases in the first intervention in Wales, all the other Covid Passport introductions appeared to be effective against the spreading of the virus. Note the significance of the coefficients, in the sense that their 95% confidence interval does not include any positive sub-interval.

**Table 2:** Estimates and 95% confidence intervals of the effect of the Covid Passport in the DiD models for the different countries and outcomes. The numbers represent incidence rates per 100000 people.

|  | Wales CP1 | Wales CP2 | Northern Ireland CP1 | Scotland CP1 |
| --- | --- | --- | --- | --- |
| Cases IR estimate | 2.2 | -7.75 | -10.1 | 7.91 |
| 95% CI cases | (-6.24,10.7) | (-13.1, -2.46) | (-18.4, -1.79) | (4.46, 11.4) |
| Admissions IR estimate | -0.144 | -0.169 | -0.269 | -0.097 |
| 95% CI admissions | (-0.248, -0.039) | (-0.308, -0.031) | (-0.385, -0.153) | (-0.219, 0.024) |

The first Covid Passport introduction in Wales was not seen effective in terms of reduction on the number of cases, compared to England, with an associated coefficient of 2.22 (95% CI -6.24,10.7). It was associated, however, with a reduction of hospitalizations, with a coefficient of -0.144 (95%CI - 0.248, -0.039). In November, the increased restriction of the Covid Passport led to a decrease in the incidence rates of both outcomes compared to England, with coefficients -7.75 (95% CI -13.1, -2.46) for incidence rate of cases and -0.169 (95% CI -0.308, -0.031) for incidence rate of hospital admissions.

Northern Ireland showed a similar result, with coefficients -10.1 (95% CI -18.4, -1.79) and -0.269 (95% CI -0.385, -0.153) for incidence rates of cases and hospital admissions respectively.

As for the number of cases in Scotland, there also seemed not to be an effect of the Covid Passport, with a coefficient of 7.91 (95% CI 4.46,11.4). Nonetheless the method indicated a significant effect on the incidence rate of hospital admissions, with a DiD coefficient of -0.097 (95% CI -0.219,0.024).

The aforementioned comparisons can be visually inspected in *Figures 1, 2* and *3*.

**Figure 1a (left) and 1b (right).**
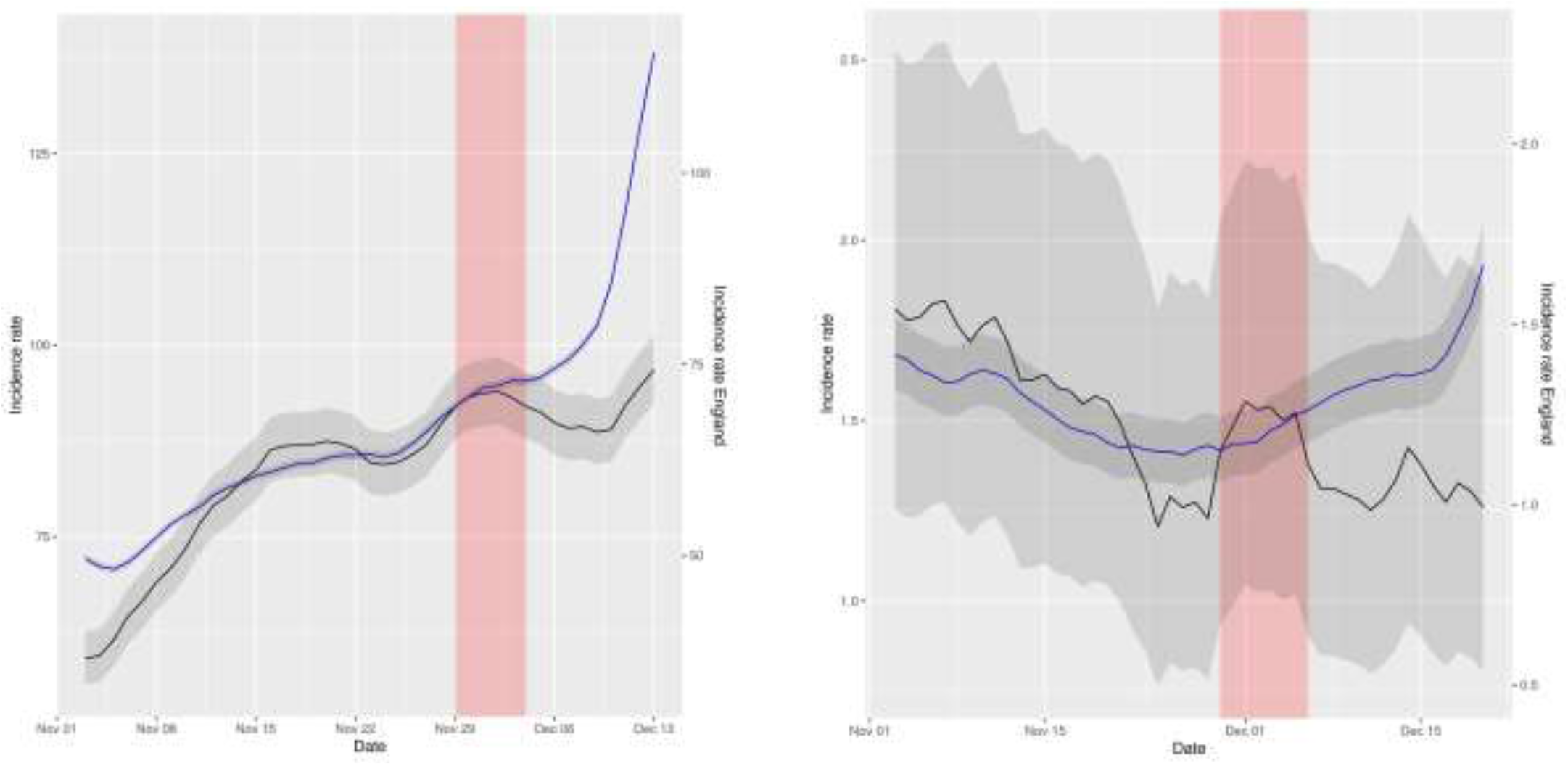
Representation of smoothed daily incidence rates of cases (left) and hospital admissions (right) in Northern Ireland (data in black) vs England (data in blue) per 10^5^ inhabitants. Data has been displaced so that both curves intersect right at the intervention time point. The red shaded area represents the neglected period post-intervention in the model due to the lag between the intervention and its effect and the grey shaded area represents the 95% confidence intervals of the calculation of incidence rates.

**Figure 2a (left) and 2b (right).**
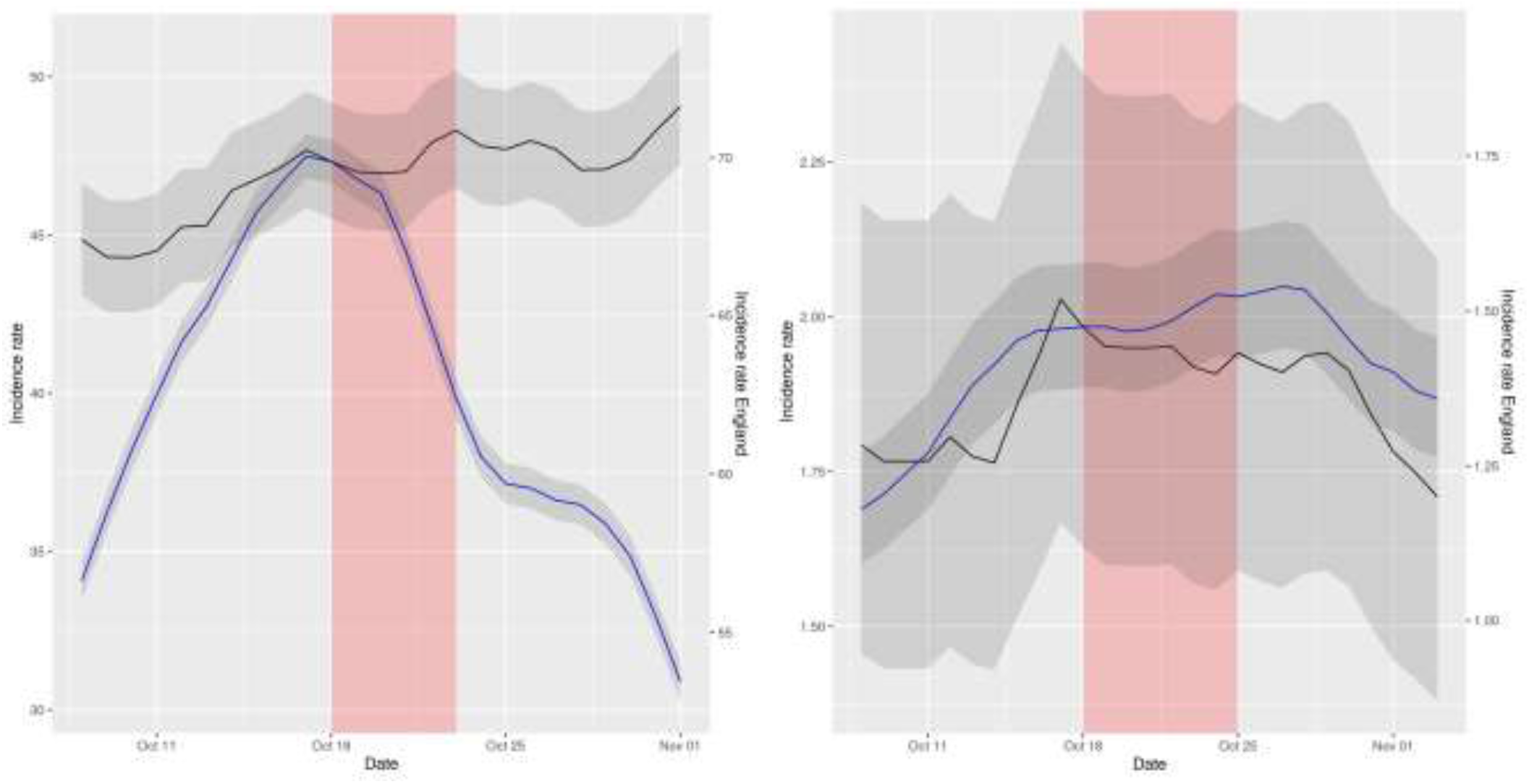
Representation of smoothed daily incidence rates of cases (left) and hospital admissions (right) in Scotland (data in black) vs England (data in blue) per 10^5^ inhabitants. Data has been displaced so that both curves intersect right at the intervention time point. The red shaded area represents the neglected period post-intervention in the model due to the lag between the intervention and its effect and the grey shaded area represents the 95% confidence intervals of the calculation of incidence rates.

**Figure 3a (left) and 3b (right).**
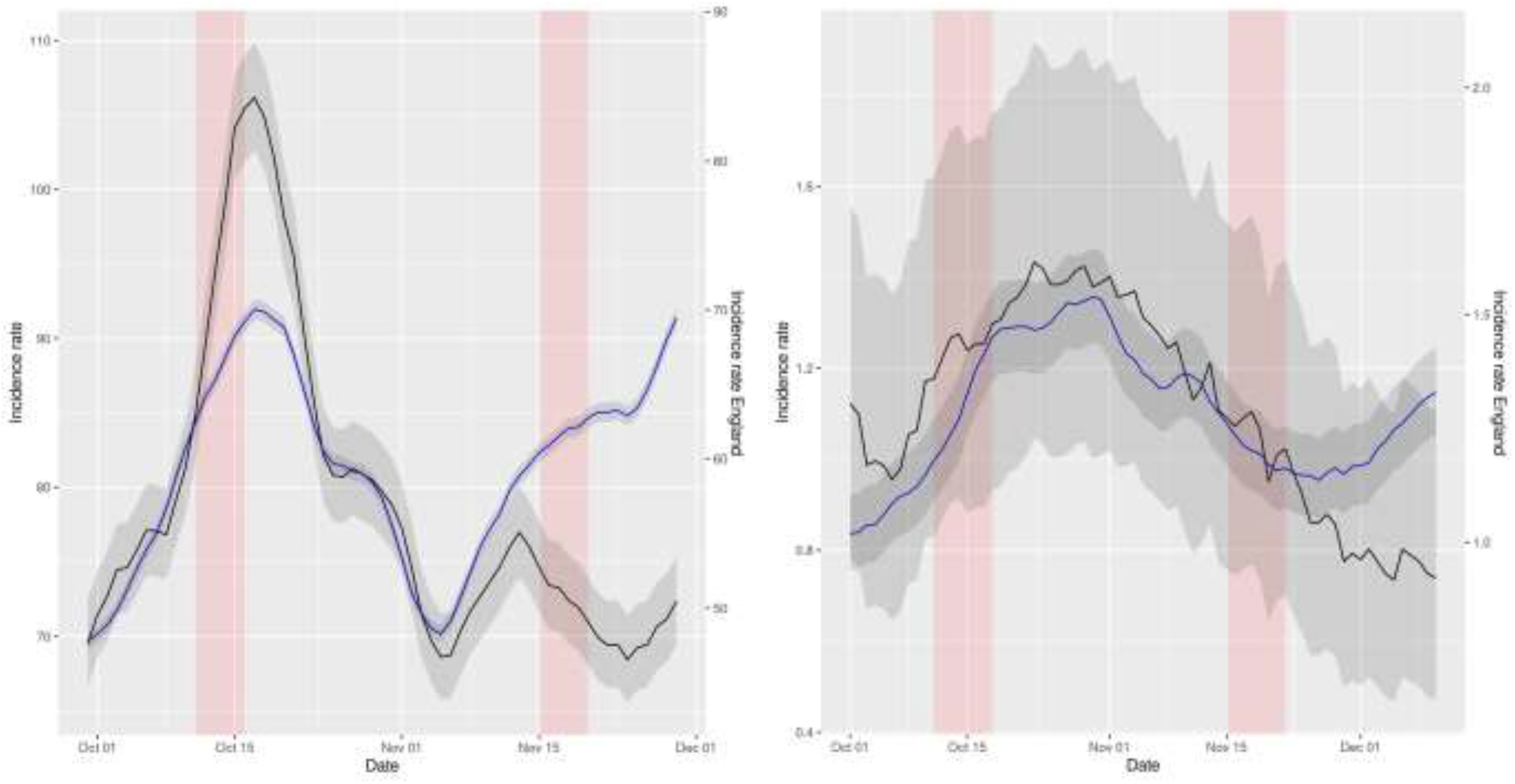
Representation of smoothed daily incidence rates of cases (left) and hospital admissions (right) in Wales (data in black) vs England (data in blue) per 10^5^ inhabitants. Data has been displaced so that both curves intersect right at the intervention time point. The red shaded area represents the neglected period post-intervention in the model due to the lag between the intervention and its effect and the grey shaded area represents the 95% confidence intervals of the calculation of incidence rates.

## Discussion

Using NBSR modelling, we found Covid Passport interventions associated with a decrease in the incidence of COVID-19 cases in all countries except England, and with a decrease in COVID-19 hospitalizations only in Scotland. ARIMA models, which control for autocorrelation of the observations, supported these findings. DiD analyses supported a causal effect of CPs to decrease incidence rates in most territories, using England as a counterfactual. However, the study of Covid Passport intervention in England itself on the 15^th^ of December 2021 shows that it was insufficient to prevent the increase in both incidence of cases and hospital admissions in the country.

This discrepancy between the effect of COVID passports in England compared to the other countries might be due to the new Omicron variant of the virus (which represented the 64% of the population of the country by that date^14^), the effect of other coexistent measures (like the mandatory use of face masks or accelerated booster vaccine campaign) or the already high uptake in vaccination. Indeed, as of 12^th^ December 2021, almost 9 in 10 individuals aged 12 and over had been vaccinated with at least one dose (42,561,679, 88.0%), more than 8 in 10 individuals aged 18 and over had been vaccinated with both doses (38,627,544, 86.9%) and more than 6 in 10 individuals aged 40 and over had received a booster or 3rd dose (18,128,105, 63.8%)^15^.

The visual difference in the NBSR plots, with England as a negative control group, reinforced the previous conclusions. Plots depicting the situation in Wales, for instance, suggested a striking effect compared to England, and in territories without geographic nor political barrier. The intervention was not associated with a reduction of hospitalizations for some countries, but even in those cases, comparing to England, the plots indicated an impact of Covid Passport on reducing the increasing trend of hospital admissions observed at the same time in the English NHS.

In the DiD analyses, we found significant effect of Covid Passport interventions for both incidence rates of cases and hospital admissions in Northern Ireland and the second half of November in Wales, compared to England, where the restriction was not into effect. The impact was not significant for the incidence rate of cases in Scotland nor October in Wales (first CP intervention), yet it was for hospital admissions. In fact, during that period the number of cases did decrease abruptly in Wales after the introduction of the Covid Passport. However, as they also decreased in England, the intervention effect was not so obvious. As for Scotland, the difference in trends pre-intervention for both groups is too acute to be able to interpret this model in a sensible way, as the assumptions for its validity are surely violated. In that sense, the DiD plots provided in the *Results* section for all regions and outcomes, compared to England, in which both trends have been superposed to better see its similarities and differences, serve as a check for the validity of this assumption. We note that this condition is arguably satisfied for all pairs except for cases in Scotland, and hence we can conclude that the reported positive effects are reliable.

These results are coherent with previous reported increased vaccine uptake after Covid Passport implementation^7,8^. Indeed, apart from the obvious restriction of mobility, the introduction of the Covid Passport and a subsequent increase in vaccine uptake could account for a lowering in both incidence of COVID-19 cases and hospitalizations. Moreover, this would explain the inefficiency observed in controlling the Omicron variant, as recent studies have reported lower effectiveness of the vaccines against infection by this variant^16,17^.

Limitations of our analyses include the aggregated nature of our data, therefore potentially limited by ecological fallacy. Time varying influential factors have possibly been controlled with DiD methods taking England as a negative control group, yet other differences between the regions might be prevalent and affect the spreading of the virus differently. Moreover, the interventions were introduced at different times and with different limitations, and the response of the population to them might have been different in different regions. An unquestionably fair comparison is thus impossible.

In conclusion, we demonstrate that the introduction of mandatory certificates was effective in decreasing cases in all countries except in England. This could be explained by differences of concomitant measures, on baseline vaccination uptake or by the emergence of the Omicron variant. Mandatory certification is only one of many policy levers to control the pandemic, and a sensible reassessment of its efficacy should be made by the competent authorities.

## Data Availability

Data was retrieved from the UK Coronavirus Dashboard (https://coronavirus.data.gov.uk/) and we
publicly host the source code at (https://github.com/KimLopezGuell/Covidpassport/blob/main/Covid_passport.r), allowing public contribution and review, and free re-use for
anyone s future research.

## Contributors

KLG is a MSc student in Mathematical Sciences at the University of Oxford. She contributed to the formal analysis, creating tables and figures, and writing the original draft of the paper. APU is a DPhil student in the Pharmacoepidemiology group at the Centre for Statistics in Medicine (CSM) (Oxford) and an MD. He helped with the conceptualisation of the project and also contributed to the formal analysis of the data and reviewed and edited the final written paper. MC is a postdoctoral scholar in the Pharmacoepidemiology group at the Centre for Statistics in Medicine (CSM) (Oxford), who also helped with the editing and review of the paper. CP is an Associate professor at the Computational Biology and Complex Systems group at the Universitat Politècnica de Catalunya. She helped with the conceptualisation and editing and review of the paper. JH is a Professor of Bioinformatics at the University of Oxford. He helped with the editing and review of the paper. DPA is a Professor of Pharmaco- and Devide Epidemiology at the Centre for Statistics in Medicine (CSM) (Oxford) and an MD. He contributed to the conceptualisation, formal analysis and editing and review of the paper.

## Ethical approval

As this research project did not involve human subject research, it was exempted from IRB approval from the participated data partners.

## Data sharing

Data was retrieved from the UK Coronavirus Dashboard (https://coronavirus.data.gov.uk/) and we publicly host the source code at (https://github.com/KimLopezGuell/Covid-passport/blob/main/Covid_passport.r), allowing public contribution and review, and free re-use for anyone’s future research.

## Transparency declaration

The lead authors affirm that this manuscript is an honest, accurate, and transparent account of the study being reported; that no important aspects of the study have been omitted; and that any discrepancies from the study as planned (and, if relevant, registered) have been explained.

## Supplementary material

### NPI dates information

| Region and intervention | Wales CP1 | Wales CP2 | Scotland CP1 | Northern Ireland CP1 | England CP1 |
| --- | --- | --- | --- | --- | --- |
| Intervention date | 2021/10/11 | 2021/11/15 | 2021/10/18 | 2021/11/29 | 2021/12/15 |
| Restrictions imposed | Covid passports – including either vaccination status or a negative Covid test within the past 48 hours – have also been required to attend nightclubs, unseated indoor events with over 500 people, unseated outdoor events with over 4,000 people, and any event with over 10,000 people. | Extended to cinemas, theatres and concert halls. | Covid passports have been required to attend nightclubs, unseated indoor events with over 500 people, unseated outdoor events with over 4,000 people, and any event with over 10,000 people. | The pass would be mandatory in the same venues as Wales and also pubs and restaurants. | Covid pass will now be mandatory for nightclubs, unseated indoor events with 500 or more attendees, unseated outdoor events with 4,000 or more attendees and any event with 10,000 or more attendees. |

### Plots for the Negative Binomial Segmented Regression models

#### Northern Ireland

**Supplementary Figure S1a (left) and S1b (right).**
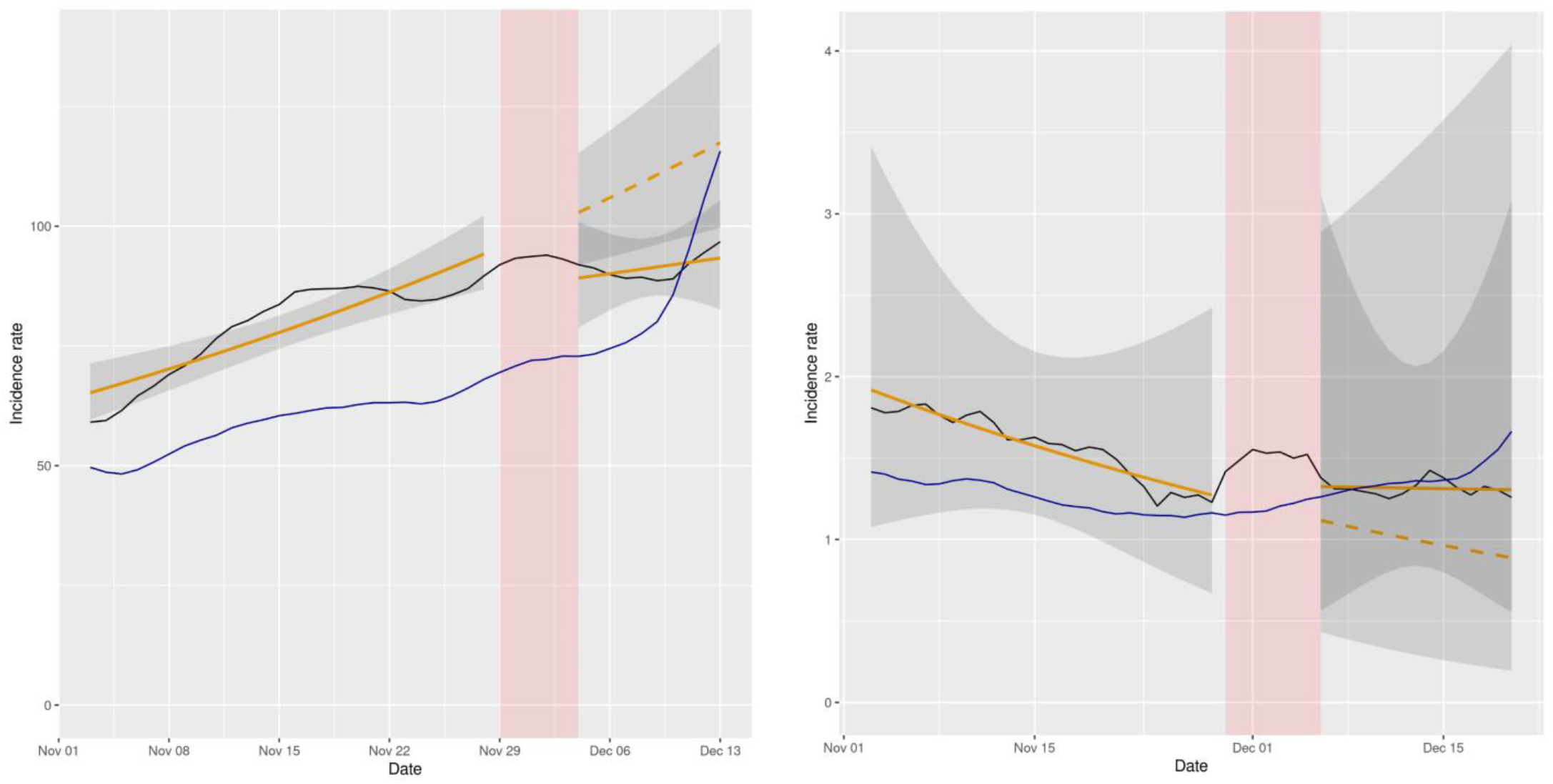
Representation of NBSR models comparing incidence rates of cases and hospital admissions in Northern Ireland (data in black, model in orange) vs England (data in blue) per 10^5^ inhabitants. Dashed lines represent the expected evolution without intervention. The red shadowed area represents the neglected period post-intervention in the model due to the lag between the intervention and its effect and the grey shaded area represents the 95% confidence intervals for the predictions.

#### Scotland

**Supplementary Figure S2a (left) and S2b (right).**
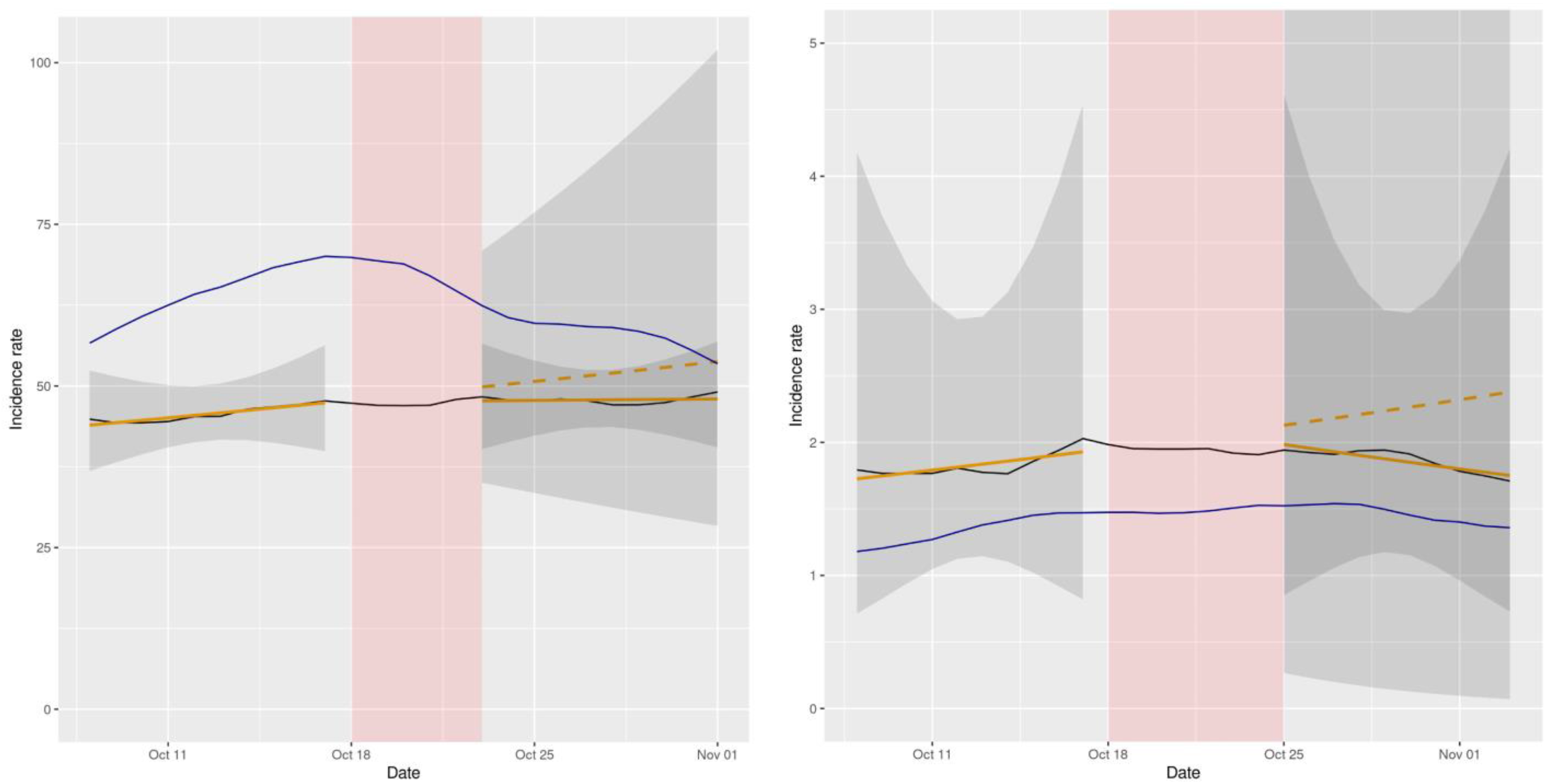
Representation of NBSR models comparing incidence rates of cases and hospital admissions in Scotland (data in black, model in orange) vs England (data in blue) per 10^5^ inhabitants. Dashed lines represent the expected evolution without intervention. The red shadowed area represents the neglected period post-intervention in the model due to the lag between the intervention and its effect and the grey shaded area represents the 95% confidence intervals for the predictions. Note: In figure S2b the y axis has been cut to make the trends more visible, as the confidence interval for the predictions post-intervention is relatively large.

#### Wales

**Figure S3a (left) and S3b (right).**
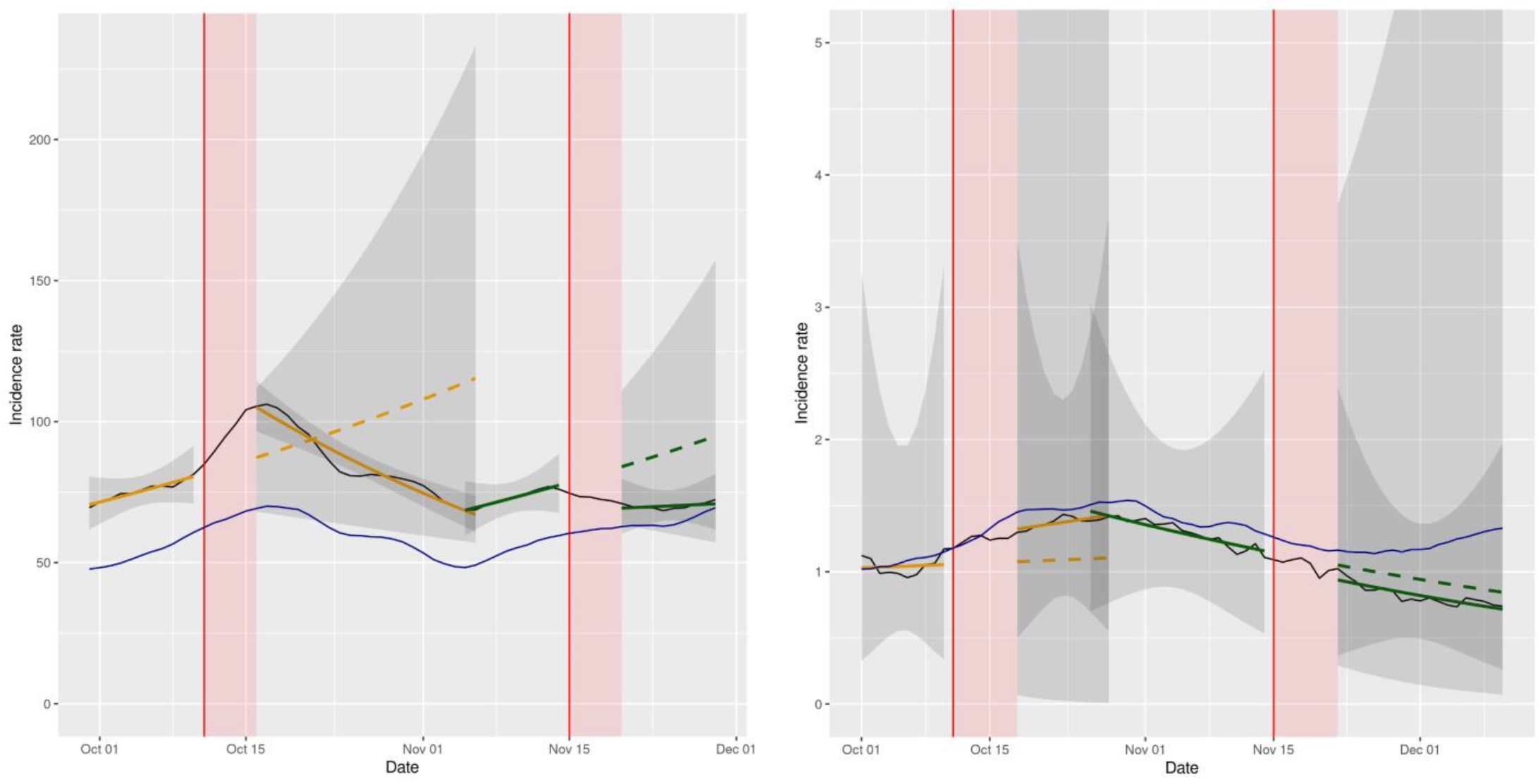
Representation of NBSR models comparing incidence rates of cases and hospital admissions in Wales (data in black, model in orange and in green) vs England (data in blue) per 10^5^ inhabitants. Dashed lines represent the expected evolution without intervention. The red shadowed area represents the neglected period post-intervention in the model due to the lag between the intervention and its effect and the grey shaded area represents the 95% confidence intervals for the predictions. Note: In figure S3b the y axis has been cut to make the trends more visible, as the confidence interval for the predictions post-intervention is relatively large.

#### England

**Figure S4a (left) and S4b (right).**
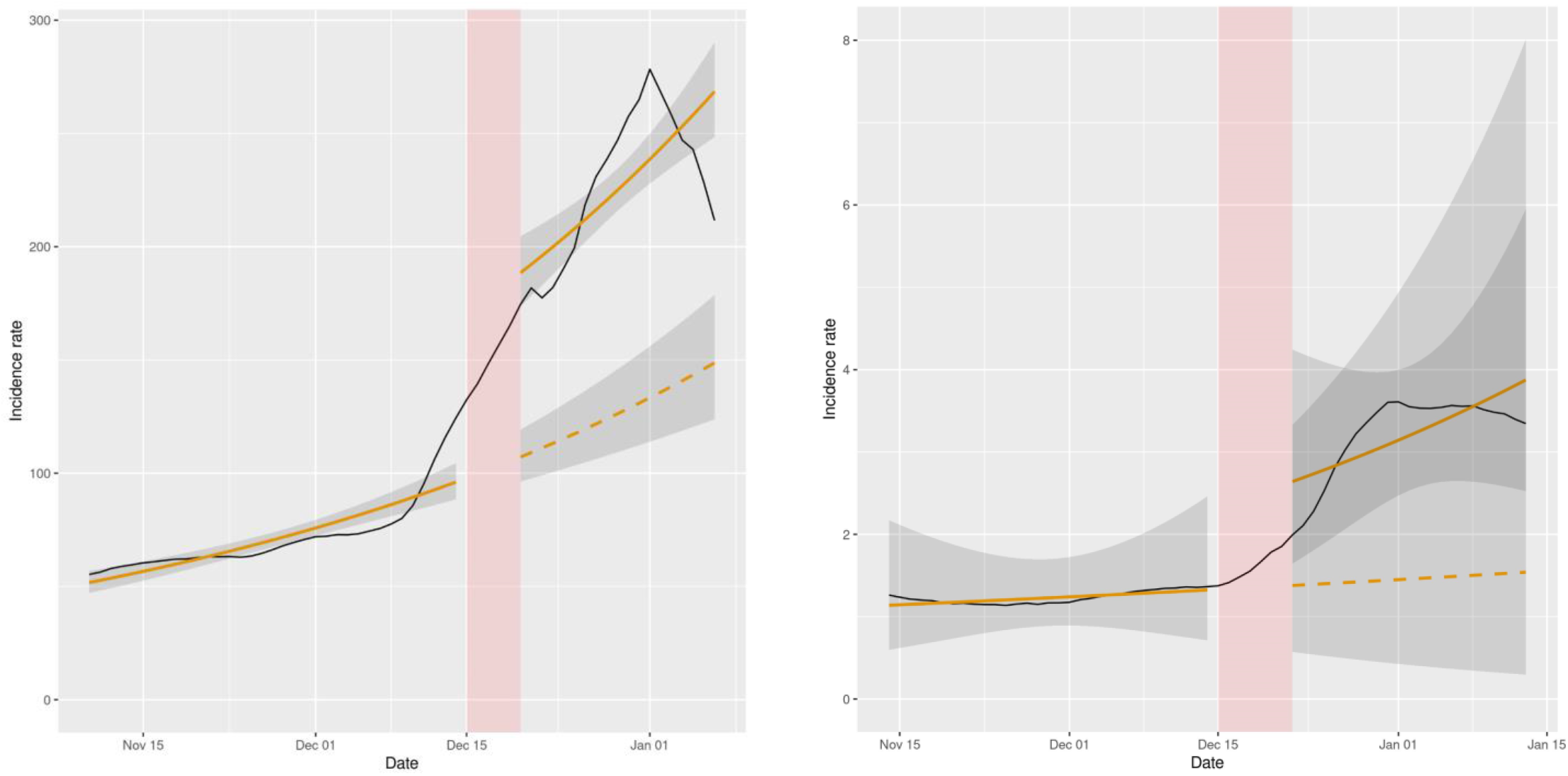
Representation of NBSR models of the incidence rates of cases and hospital admissions in England (data in black, model in orange) per 10^5^ inhabitants. Dashed lines represent the expected evolution without intervention. The red shadowed area represents the neglected period post-intervention in the model due to the lag between the intervention and its effect and the grey shaded area represents the 95% confidence intervals for the predictions.

